# Association of genetic liability to smoking initiation with e-cigarette use in young adults

**DOI:** 10.1101/2020.06.10.20127464

**Authors:** Jasmine N Khouja, Robyn E Wootton, Amy E Taylor, George Davey Smith, Marcus R Munafò

## Abstract

**Background:** Smoking and e-cigarette use are strongly associated, but it is currently unclear whether this association is causal, or due to shared factors that influence both behaviours such as a shared genetic liability. The aim of this study was to investigate whether polygenic risk scores (PRS) for smoking initiation are associated with ever use of e-cigarettes.

**Methods and Findings:** PRS of smoking initiation were calculated for young adults (aged 23 to 26 years) of European ancestry in the Avon Longitudinal Study of Parents and Children using the GWAS & Sequencing Consortium of Alcohol and Nicotine use (GSCAN) summary statistics. Five thresholds ranging from 5×10^−8^ to 0.5 were used to calculate five PRS for each individual. Using logistic regression, we investigated the association between smoking initiation PRS and both self-reported smoking initiation and self-reported e-cigarette use, as well as a number of negative control outcomes (socioeconomic position at birth, externalising disorders in childhood and risk-taking in young adulthood). We observed positive associations of similar magnitude between smoking initiation PRS and both smoking initiation (OR = 1.29, 95% CI 1.19 to 1.39) and ever e-cigarette use (OR = 1.24, 95% CI 1.14 to 1.34) by the age of 24 years. At lower *p*-value thresholds, we observed an association between smoking initiation PRS and ever e-cigarette use among never smokers. We also found evidence of associations between smoking initiation PRS and some negative control outcomes, particularly when less stringent *p*-value thresholds were used but also at the strictest threshold (e.g., gambling, number of sexual partners, conduct disorder at 7 years, and parental socioeconomic position at birth).

**Conclusions:** Our results indicate that there may be a shared genetic aetiology between smoking and e-cigarette use, and also with socioeconomic position, externalising disorders in childhood, and risky behaviour more generally. Taken together, this indicates that there may be a common genetic vulnerability to both smoking and e-cigarette use, which may reflect a broad risk-taking phenotype.

## Introduction

There are an estimated 3.6 million electronic cigarette (e-cigarette) users in Great Britain [1] and evidence is growing that e-cigarettes are effective in helping smokers quit [2, 3]. The use of e-cigarettes for smoking cessation is common among young adults in the UK (Khouja, Taylor, & Munafò, 2019); therefore, it would be logical to assume that smoking causally influences e-cigarette use in this population. However, some studies have shown an association between e-cigarette use and subsequent smoking among non-smokers, which suggests the possibility that e-cigarette use may also act as a gateway to smoking (sometimes referred to as the gateway hypothesis), particularly among adolescents. A recent meta-analysis found that for young people aged 30 years or younger there is a strong and consistent positive association between e-cigarette use among never smokers and later smoking, but that there is currently insufficient evidence to conclude that this association is causal [4]. Understanding more about the nature of the association between smoking and e-cigarette use, particularly in young adulthood, is vital to inform tobacco control policies that aim to prevent youth smoking initiation by restricting access to e-cigarettes. Specifically, it is important to understand whether the association found among young adults is causal, or due to other factors that influence both smoking and e-cigarette use independently.

For example, there is some evidence for a shared genetic liability to both smoking and e-cigarette use [5]. This could indicate a causal relationship in that genetic variants influence smoking which then increases the probability of vaping (i.e., vertical pleiotropy), or it could be due to genetic variants that influence a phenotype which consequently influences both behaviours (i.e., horizontal pleiotropy) [6]. One biologically plausible explanation for a genetic link between smoking and e-cigarette use is that they are both influenced by the same genetic variants that influence an individual’s response to nicotine or their nicotine metabolism. However, evidence suggests that some of the genetic influence on smoking initiation is mediated by personality traits, such as risk-taking and impulsivity, that influence (among other things) smoking uptake [7]. Allegrini and colleagues [5] suggest that a genetic link between smoking and e-cigarette use may reflect these personality traits (i.e., a genetic liability to take risks may influence an individual’s likelihood of initiating smoking and vaping).

Using genetic variants, we can explore whether smoking is associated with e-cigarette use, and which factors or mechanisms may influence the association. Ideally, we would explore the genetic overlap between smoking and e-cigarette use by comparing the genetic variants identified in genome wide association studies (GWAS) of each behaviour, but at present there are no large, well-powered GWAS of e-cigarette use. However, a GWAS of various smoking behaviours has recently been published [8], which identified 378 single nucleotide polymorphisms (SNPs) associated with smoking initiation. Using these SNPs, smoking initiation polygenic risk scores (PRS) can be created and associations between these PRS and a range of outcomes examined.

Causality cannot be inferred from such analyses, but negative control outcomes can be used to inform the overall evaluation of whether an association is causal via a hypothesised route. Negative controls are outcomes which are not plausibly caused by the exposure – for example, smoking is associated with risk of dying by suicide (which is biologically plausible), but equally strongly associated with risk of dying by homicide (which is not), casting doubt on the causal nature of the former association [9]. Triangulating evidence from outcomes where a simple biological pathway from smoking to the outcome is implausible (e.g., gambling), or impossible (e.g., externalising behaviour or socioeconomic position [SEP] in childhood, before smoking has occurred) can aid consideration of whether any association between genetic liability to smoking and e-cigarette use is likely to be due to a biological pathway from smoking to e-cigarette use (i.e., vertical pleiotropy), or due to the genetic liability to smoking capturing a broader, risk-taking phenotype (i.e., horizontal pleiotropy). Alternatively, triangulation could aid consideration of whether an association is due to a shared genetic predisposition between parents and offspring; where parents share their offspring’s smoking initiation predisposition and consequently expose their offspring to cigarette smoke in utero or in childhood, an apparent effect of a child’s own genetic variants may be a result of their pre-natal or post-natal environment due to their parents’ genetic variants. If associations are only found between smoking initiation PRS and e-cigarette use, but not negative control outcomes, this would strengthen the vertical pleiotropy interpretation; however, if an association is also found with negative control outcomes, this would indicate that horizontal pleiotropy is occurring or that shared parent-offspring genetic predisposition may be confounding the association.

Additionally, using varying *p*-value thresholds to create PRS could help to identify the presence of horizontal pleiotropy. Calculating PRS at less strict *p*-value thresholds than the standard genome wide significant threshold increases the percentage variance in the phenotype explained by the score, and thus increases power to detect an association. However, using less stringent thresholds will also tend to increase the likelihood of including genetic variants which are related to other factors, making the PRS less specific to the exposure of interest (and may eventually result in PRS which explain less variance in the exposure). The more SNPs included in a PRS, the less likely it is that the effect of each variant on the trait of interest is proportional to the effect of the trait of interest on the exposure, and the more likely it is proportional to the effects on other (horizontally pleiotropic) traits [10], increasing the likelihood that any associations found between the PRS and an outcome could be due to horizontal pleiotropy. Triangulating evidence from a variety of thresholds and a variety of outcomes may provide a clearer picture of the true association; associations observed when more stringent PRS thresholds are used could be due to a causal effect of smoking, while associations observed only at less stringent thresholds among negative control outcomes may indicate horizontal pleiotropy.

We aimed to investigate whether smoking initiation PRS are associated with ever use of e-cigarettes in young adulthood. We also aimed to explore any associations with outcomes that are not plausibly biologically related (e.g., gambling) or that precede smoking (e.g., hyperactivity in childhood), to determine whether the association between smoking and e-cigarette use could reflect a broader risk-taking phenotype captured by the smoking initiation PRS.

## Methods

### Data Sources

#### GSCAN

The GWAS & Sequencing Consortium of Alcohol and Nicotine use (GSCAN) report summary level statistics from a GWAS of smoking initiation [8]. This GWAS was based on 1,232,091 participants from 29 cohorts. Summary statistics, with the Avon Longitudinal Study of Parents and Children (ALSPAC; N = 11,345) removed, were obtained through correspondence with GSCAN to eliminate data overlap with the target sample. Due to data sharing restrictions, 23andMe were also excluded from this data (N = 599,289) leaving a total sample size of 621,457. Smoking initiation was defined as ever being a regular smoker. The exact definition varied across the cohorts included in the GWAS, with 3 different definitions: 1) Have you smoked over 100 cigarettes over the course of your life? 2) Have you ever smoked every day for at least a month? 3) Have you ever smoked regularly?

#### ALSPAC

The target sample consisted of participants from ALSPAC [11, 12]. This study recruited pregnant women residing in Avon, UK with expected dates of delivery 1st April 1991 to 31st December 1992. The phases of enrolment are described in detail in the cohort profile paper and its update [13]. A total of 15,454 mothers were recruited, resulting in 15,589 foetuses. Of these, 14,901 were alive at 1 year of age. The study website contains details of all the data that is available through a fully searchable data dictionary and variable search tool (http://www.bristol.ac.uk/alspac/researchers/our-data/). Genetic data was available for 9,085 young adults and, after samples which did not pass quality control were removed, PRS were created for 7,859 unrelated individuals of European ancestry. Of these individuals, 2,905 also had data for our main outcome at 24 years regarding their vaping behaviour. ALSPAC study data from 22 years onwards were collected and managed using REDCap electronic data capture tools hosted at the University of Bristol [14]. Sample sizes varied by outcome due to restrictions (e.g., restricting to never smokers) and differing timepoints of measurement (i.e., missing data). Ethics approval for the study was obtained from the ALSPAC Ethics and Law Committee and the Local Research Ethics Committees. Consent for biological samples has been collected in accordance with the Human Tissue Act (2004). Informed consent for the use of data collected via questionnaires and clinics was obtained from participants following the recommendations of the ALSPAC Ethics and Law Committee at the time.

#### Polygenic Risk Scores

Summary data from GSCAN (excluding ALSPAC and 23andMe, N = 621,457) were used to select SNPs associated with smoking initiation. Betas were converted to log odds ratios. Each participant was given a score which indicated the average number of risk alleles (0, 1 or 2 effect alleles) they possessed for the selected SNPs. Scores were weighted (i.e., multiplied) by the regression coefficients from the summary statistics (with ALSPAC and 23andMe removed), then standardised by transforming to z-scores. Five *p*-value thresholds (5×10^−8^, 0.0005, 0.005, 0.05, 0.5) were used to determine five groups of SNPs to be included in five different PRS for each participant. PLINK was used to determine PRS at the *p* < 5×10^−8^ threshold using the SNPs which met the genome wide significance threshold in the GSCAN GWAS of smoking initiation [8]. PRSice software was used to calculate the PRS at all other thresholds [15]. The data acquired from GSCAN was pruned for SNPs with a Minor Allele Frequency (MAF) > 0.001 where at least 10% of the maximum sample size had SNP data available in at least three of the consortium studies. SNPs were clumped to ensure low linkage disequilibrium (r^2^ < 0.1).

#### Outcomes

Detailed information regarding the phenotype data including the questions and answer options provided in the questionnaires are available in Supplementary Material (Table S1).

#### E-cigarette use

At 24 years (between 2016 and 2017), outcome data was collected via questionnaire on whether participants had ever vaped. Ever use was defined as ever having used/vaped an e-cigarette or other vaping device.

#### Smoking

Self-reported smoking initiation and ever smoking were included as positive control outcomes (i.e., outcomes for which an association with the exposure is expected). Smoking initiation by 24 years was defined as having smoked 100 or more cigarettes in their lifetime. Ever smoking by 24 years was defined as having ever smoked a whole cigarette (including roll-ups).

#### Negative controls

Four negative control outcomes at age 23 and 24 were included in the analysis: high number of sexual partners, having been in trouble with the law, ever gambling, and enjoying taking risks. These were selected on the basis of being related to broad risk-taking behaviour, but where a causal pathway from smoking was not considered biologically plausible. Three negative control outcomes at age 7 were included: hyperactivity, conduct disorder (CD) and oppositional defiant disorder (ODD). These externalising disorders are indicators of impulsivity and were selected on the basis that few (if any) children at this age have smoked, ruling out a causal pathway from their own smoking to these outcomes. Parental SEP, which was measured at birth, was also included in the analysis. This outcome was based on highest occupation of both parents at birth (preceding smoking) and was selected on the basis that it could not possibly be caused by a young person’s own smoking. Further information regarding the negative controls can be found in the Supplementary Material.

#### Statistical Analyses

All analyses were carried out in STATA 15.1 [16] using logistic regression and were adjusted for age, sex and the first 10 principal components. We assessed the association between smoking initiation PRS and (i) ever e-cigarette use by age 24 among the full sample and those who had never smoked, (ii) regular e-cigarette use at age 24, (iii) smoking initiation, and (iv) negative control outcomes (risk taking behaviours, externalising disorders, and SEP).

## Results

A total of 378 SNPs were identified as genome wide significant in the GSCAN GWAS of smoking initiation [8], 356 of which were available in ALSPAC. Nine SNPs were removed at the clumping stage, leaving 347 SNPs included in the most stringent PRS (*p*-value threshold *p* < 5×10^−8^). The number of SNPs included in each PRS at the less stringent thresholds are shown in Table S2. Of note, PRS calculated at these less stringent thresholds were based on the significance level reported in the restricted sample (excluding ALSPAC and 23andMe) summary data.

Table 1 shows the characteristics of the sample; 878 (30%) young adults were self-reported ever e-cigarette users by 24 years and 1,695 (64%) were self-reported ever smokers. Of those who had ever vaped, 95% (n=830) had ever smoked at least one whole cigarette and 71% (n=616) had smoked 100 or more cigarettes. Less than 1% of the sample had used an e-cigarette prior to smoking. Self-reported smoking and e-cigarette use were associated with lower parental SEP and having externalising disorders in childhood (Table S3). Self-reported smoking and e-cigarette use were also associated with increased odds of engaging in risk-taking behaviours (Table S3).

**Table 1.** Characteristics of young adults in ALSPAC

| Characteristic | N (%) |
| --- | --- |
| Ever used an e-cigarette by 24 (used once or more) | 878 (30%) |
| Regularly used an e-cigarette at 24 (used at least once a month) | 150 (5%) |
| Ever smoked by 24 (1 cigarette or more) | 1,695 (64%) |
| Initiated smoking by 24 (100 cigarettes or more) | 972 (33%) |
| Ever used an e-cigarette but not initiated smoking by 24 | 262 (13%) |
| High number of sexual partners at 23* | 647 (25%) |
| Been in trouble with the law since 23rd birthday | 69 (2%) |
| Enjoys taking risks at 24 | 1,618 (55%) |
| Ever gambled at 24 | 2,156 (74%) |
| Hyperactivity at 7 | 2,219 (42%) |
| Conduct disorder at 7 | 1,199 (22%) |
| Oppositional defiant disorder at 7 | 1,868 (35%) |
| Parental SEP (manual) | 1,068 (27%) |
|  | Mean (SD) |
| Age in months at 24 year questionnaire | 298 (6) |

*Note:* Sample sizes varied by characteristic due to differing timepoints of measurement (i.e., missing data). \* 11 or more sexual partners.

### Smoking Initiation PRS and Self-Reported Smoking

We observed positive associations between smoking initiation PRS and ever smoking (having smoked at least 1 cigarette in a lifetime) by the age of 24 years (*p* < 5×10^−8^ threshold OR (OR_10-8_) = 1.25, 95% CI 1.16 to 1.35) and smoking initiation (having smoked at least 100 cigarettes in a lifetime) by the age of 24 years (OR_10-8_ = 1.29, 95% CI 1.19 to 1.39). We found strong associations between smoking initiation PRS and self-reported smoking measures at all *p*-value thresholds (Table 2).

**Table 2.** Associations between polygenic risk scores for smoking initiation with ever e-cigarette use, ever smoking and smoking initiation.

| **Outcome** | *p*-value threshold | n | OR | 95% CI | *p* |
| --- | --- | --- | --- | --- | --- |
| **Ever e-cigarette use by 24** |  | 2,894 |  |  |  |
|  | 5x10<sup>-8</sup> |  | 1.24 | 1.14, 1.34 | <0.001 |
|  | 0.0005 |  | 1.27 | 1.17, 1.38 | <0.001 |
|  | 0.005 |  | 1.36 | 1.26, 1.48 | <0.001 |
|  | 0.05 |  | 1.39 | 1.28, 1.51 | <0.001 |
|  | 0.5 |  | 1.39 | 1.28, 1.51 | <0.001 |
| **Regular e-cigarette use at 24 (at least once a month)** |  | 2,894 |  |  |  |
|  | 5x10<sup>-8</sup> |  | 1.18 | 1.00, 1.40 | 0.049 |
|  | 0.0005 |  | 1.22 | 1.03, 1.44 | 0.019 |
|  | 0.005 |  | 1.22 | 1.04, 1.44 | 0.017 |
|  | 0.05 |  | 1.18 | 1.00, 1.39 | 0.051 |
|  | 0.5 |  | 1.22 | 1.04, 1.44 | 0.018 |
| **Ever smoking by 24 (1 cigarette or more)** |  | 2,931 |  |  |  |
|  | 5x10<sup>-8</sup> |  | 1.25 | 1.16, 1.35 | <0.001 |
|  | 0.0005 |  | 1.27 | 1.17, 1.38 | <0.001 |
|  | 0.005 |  | 1.32 | 1.22, 1.43 | <0.001 |
|  | 0.05 |  | 1.33 | 1.23, 1.44 | <0.001 |
|  | 0.5 |  | 1.34 | 1.24, 1.44 | <0.001 |
| **Smoking initiation (100 cigarettes or more) by 24** |  | 2,925 |  |  |  |
|  | 5x10<sup>-8</sup> |  | 1.29 | 1.19, 1.39 | <0.001 |
|  | 0.0005 |  | 1.38 | 1.27, 1.49 | <0.001 |
|  | 0.005 |  | 1.46 | 1.34, 1.58 | <0.001 |
|  | 0.05 |  | 1.49 | 1.37, 1.61 | <0.001 |
|  | 0.5 |  | 1.49 | 1.37, 1.39 | <0.001 |
| **Ever e-cigarette use by 24 among never smokers (<100 cigarettes)** |  | 1,937 |  |  |  |
|  | 5x10<sup>-8</sup> |  | 1.10 | 0.97, 1.26 | 0.150 |
|  | 0.0005 |  | 1.05 | 0.92, 1.20 | 0.464 |
|  | 0.005 |  | 1.12 | 0.98, 1.28 | 0.087 |
|  | 0.05 |  | 1.15 | 1.00, 1.31 | 0.046 |
|  | 0.5 |  | 1.18 | 1.04, 1.35 | 0.012 |

Note: Ever smoking and smoking initiation models were included as positive controls. Analyses were adjusted for age, sex and principal components 1-10.

### Smoking Initiation PRS and Self-Reported E-cigarette Use

We observed positive associations between smoking initiation PRS and self-reported ever use of e-cigarettes by the age of 24 years (OR_10-8_ = 1.24, 95% CI 1.14 to 1.34) and self-reported regular (at least once a month) e-cigarette use at 24 years (OR_10-8_ = 1.18, 95% CI 1.00 to 1.40). We observed these associations at all *p*-value thresholds (Table 2). Among those who had never initiated smoking (i.e., smoked < 100 cigarettes in their lifetime), we found no clear evidence for an association between smoking initiation PRS and ever e-cigarette use at the most stringent *p*-value thresholds. However, we found evidence of a positive association with PRS calculated using less stringent thresholds (*p* < 0.5 threshold OR = 1.18, 95% CI 1.04 to 1.35; Table 2). We found similar patterns of association among those who had never smoked any cigarettes (Table S4).

### Smoking Initiation PRS and Negative Controls

We observed a positive association between smoking initiation PRS and high number of sexual partners by 23 years (OR_10-8_ = 1.15, 95% CI 1.05 to 1.26) and having ever gambled by 24 years (OR_10-8_ = 1.12, 95% CI 1.03 to 1.22) at all *p*-value thresholds (Table 3). We found some evidence of a positive association between smoking initiation PRS and enjoying taking risks at 24 years (OR_0.005_ = 1.11, 95% CI 1.03 to 1.19), but this was less clear at the more stringent thresholds (Table 3). There was no clear evidence of an association between smoking initiation PRS and having been in trouble with the law since their 23^rd^ birthday (Table 3).

**Table 3.** Associations between polygenic risk scores for smoking initiation with negative controls of risky behaviour.

| Outcome | *p*-value threshold | n | OR | 95% CI | *p* |
| --- | --- | --- | --- | --- | --- |
| **Number of sexual partners by 23\*** |  | 2,505 |  |  |  |
|  | 5x10<sup>-8</sup> |  | 1.15 | 1.05, 1.26 | 0.003 |
|  | 0.0005 |  | 1.12 | 1.02, 1.23 | 0.019 |
|  | 0.005 |  | 1.18 | 1.08, 1.29 | <0.001 |
|  | 0.05 |  | 1.25 | 1.14, 1.37 | <0.001 |
|  | 0.5 |  | 1.30 | 1.19, 1.43 | <0.001 |
| **Been in trouble with the law since 23<sup>rd</sup> birthday** |  | 2,928 |  |  |  |
|  | 5x10<sup>-8</sup> |  | 1.00 | 0.79, 1.28 | 0.988 |
|  | 0.0005 |  | 1.12 | 0.88, 1.43 | 0.352 |
|  | 0.005 |  | 1.11 | 0.87, 1.41 | 0.407 |
|  | 0.05 |  | 1.04 | 0.82, 1.33 | 0.745 |
|  | 0.5 |  | 0.90 | 0.71, 1.15 | 0.394 |
| **Enjoys taking risks at 24** |  | 2,932 |  |  |  |
|  | 5x10<sup>-8</sup> |  | 1.06 | 0.98, 1.14 | 0.154 |
|  | 0.0005 |  | 1.05 | 0.98, 1.14 | 0.163 |
|  | 0.005 |  | 1.11 | 1.03, 1.19 | 0.005 |
|  | 0.05 |  | 1.09 | 1.01, 1.17 | 0.029 |
|  | 0.5 |  | 1.08 | 1.01, 1.16 | 0.033 |
| **Ever gambled by 24** |  | 2,899 |  |  |  |
|  | 5x10<sup>-8</sup> |  | 1.12 | 1.03, 1.22 | 0.008 |
|  | 0.0005 |  | 1.16 | 1.07, 1.26 | 0.001 |
|  | 0.005 |  | 1.16 | 1.06, 1.26 | 0.001 |
|  | 0.05 |  | 1.20 | 1.10, 1.30 | <0.001 |
|  | 0.5 |  | 1.15 | 1.06, 1.25 | 0.001 |

Note: Number of sexual partners, trouble with the law, enjoying risk taking and gambling models were included as negative controls. Analyses were adjusted for age, sex and principal components 1-10. \*Low (<11) vs. high (11 or more) number of sexual partners.

We found evidence of a positive association between smoking initiation PRS and hyperactivity at 7 years (OR_0.0005_ = 1.10, 95% CI 1.04 to 1.16) but not at the most stringent threshold (Table 4). There was also a positive association with CD at 7 years (OR_10-8_ = 1.10, 95% CI 1.03 to 1.17) at all thresholds (Table 4). There was some evidence of a positive association between PRS and ODD specifically at the 0.0005 threshold (OR_0.0005_ = 1.08, 95% CI 1.02 to 1.14). We also found a positive association with lower parental SEP (OR_10-8_ = 1.08, 95% CI 1.01 to 1.16) at all thresholds (Table 5).

**Table 4.** Associations between polygenic risk scores for smoking initiation with negative controls of externalising disorders in childhood.

| Outcome | *p*-value threshold | n | OR | 95% CI | *p* |
| --- | --- | --- | --- | --- | --- |
| **Hyperactivity at 7** |  | 5,227 |  |  |  |
| | $5 \times 10^{-8}$ | | 1.02 | 0.96, 1.08 | 0.511 |
|  | 0.0005 |  | 1.10 | 1.04, 1.16 | 0.001 |
|  | 0.005 |  | 1.14 | 1.08, 1.20 | <0.001 |
|  | 0.05 |  | 1.14 | 1.08, 1.21 | <0.001 |
|  | 0.5 |  | 1.15 | 1.08, 1.21 | <0.001 |
| **Conduct disorder at 7** |  | 5,334 |  |  |  |
| | $5 \times 10^{-8}$ | | 1.10 | 1.03, 1.17 | 0.004 |
|  | 0.0005 |  | 1.11 | 1.04, 1.19 | 0.001 |
|  | 0.005 |  | 1.11 | 1.04, 1.18 | 0.002 |
|  | 0.05 |  | 1.08 | 1.01, 1.15 | 0.021 |
|  | 0.5 |  | 1.08 | 1.01, 1.15 | 0.017 |
| **Oppositional defiant disorder at 7** |  | 5,325 |  |  |  |
| | $5 \times 10^{-8}$ | | 1.02 | 0.96, 1.08 | 0.496 |
|  | 0.0005 |  | 1.08 | 1.02, 1.14 | 0.013 |
|  | 0.005 |  | 1.04 | 0.98, 1.10 | 0.200 |
|  | 0.05 |  | 1.04 | 0.98, 1.10 | 0.173 |
|  | 0.5 |  | 1.02 | 0.96, 1.08 | 0.529 |

Note: Hyperactivity and conduct disorder were assessed using the strengths and difficulties questionnaire (SDQ) and oppositional defiant disorder was assessed using the development and wellbeing assessment (DAWBA). All variables were recoded into binary outcomes (no disorder/symptoms versus borderline/disorder/symptoms).

**Table 5.** Associations between polygenic risk scores for smoking initiation with negative controls of socioeconomic indicators.

| Outcome | *p*-value threshold | n | OR | 95% CI | *p* |
| --- | --- | --- | --- | --- | --- |
| **Parental SEP (manual)** |  | 6,702 |  |  |  |
|  | 5x10<sup>-8</sup> |  | 1.08 | 1.01, 1.16 | 0.017 |
|  | 0.0005 |  | 1.13 | 1.06, 1.21 | <0.001 |
|  | 0.005 |  | 1.16 | 1.09, 1.24 | <0.001 |
|  | 0.05 |  | 1.11 | 1.03, 1.18 | 0.003 |
|  | 0.5 |  | 1.13 | 1.05, 1.20 | <0.001 |

Note: SEP = Socioeconomic position. Parental SEP was based on the higher of the mother or partner's occupational social class using the Office of Population Censuses and Surveys (OPCS) occupation codes.

## Discussion

In contrast to the results of Allegrini and colleagues [5], smoking initiation PRS were strongly associated with ever e-cigarette use by 24 years. As expected, we observed an association of smoking initiation PRS and both ever smoking and smoking initiation. It was notable that the associations of the smoking initiation PRS and both smoking and e-cigarette use were of similar magnitude.

The association between smoking initiation PRS and e-cigarette use could be explained by smoking causally influencing e-cigarette use. This hypothesis is supported by observational evidence; use of e-cigarettes for smoking cessation is common among both young adults in the UK [17] and adults in Great Britain [18]. However, the associations observed among the restricted analysis and between the negative control outcomes suggest there may be other factors at play – there may be shared genetic risk factors that influence both behaviours. Among never smokers, we found weak evidence of an association between smoking initiation PRS and e-cigarette use, which suggests that the e-cigarette use is not simply caused by smoking (which has not occurred in these cases) but that there is a shared genetic aetiology influencing both behaviours. Hence, what appears to be a gateway between e-cigarette use and smoking in previous studies could actually be a shared genetic liability, and the order of use is coincidental or due to other factors such as perceived risk or mis-reporting of smoking status [19].

Alternatively, the smoking initiation PRS may be capturing much more than just smoking or nicotine use. Using less stringent *p*-value thresholds to create PRS increases the percentage variance in the phenotype explained by the score, and therefore the power to detect an association up to a point; using less stringent thresholds also increases the likelihood of capturing SNPs which are related to other factors, which adds noise and eventually results in less specific PRS that explain less variance in the exposure and more variance in other (horizontally pleiotropic) effects. Increasing magnitudes of association with PRS and negative controls at less stringent *p*-value thresholds suggests that the smoking initiation PRS is capturing, at least in part, a broad phenotype which is not entirely specific to smoking/nicotine. Although weaker associations were observed between risk-taking factors and PRS for smoking initiation compared to e-cigarette use and smoking, the associations are still relatively strong and consistent. Recent observational evidence also indicated a strong association between e-cigarette use and smoking prior to adjusting for risk taking behaviours and other shared risk factors, but showed no clear evidence of an association after adjusting for risk taking behaviours and other shared risk factors [20]. We also found an association between the smoking initiation PRS and externalising disorders in childhood (7 years) which precedes the age at which cigarettes are first smoked in the vast majority of cases in this cohort (>99%), and therefore cannot be a causal effect of own smoking. However, this association could potentially be due to causal *in utero* effects of maternal smoking in pregnancy or maternal smoking in childhood, since maternal and offspring genotype will be correlated. Nevertheless, combined with evidence that liability to ADHD increases the likelihood of smoking initiation and vice versa [21], our results suggest the possibility that the smoking initiation PRS is capturing a broad impulsivity phenotype. The association observed between PRS for smoking initiation and parental SEP also suggests the PRS could be capturing sociodemographic factors as well as smoking.

The associations observed here may also have implications for the use of smoking initiation PRS in Mendelian randomisation (MR) analysis. This method is often implemented to provide unconfounded causal estimates, as long as the assumptions of MR hold [22]. One assumption is that the genetic instrument (e.g., smoking initiation PRS) is not associated with any confounders (e.g., risk taking, childhood externalising disorders, SEP). The association we observed between smoking initiation PRS and negative control outcomes, even when restricted to only genome wide significant SNPs, indicates that smoking initiation PRS may not be a valid instrument to use in MR to investigate the causal effects of smoking initiation. This emphasises the importance of using pleiotropy robust methods (e.g., MR Egger). The InSIDE (Instrument Strength Independent of Direct Effect) assumption requires that SNP-exposure effects (e.g., the effect of smoking initiation SNPs on smoking initiation) should not be correlated with horizontal pleiotropic effects (e.g., the effect of smoking initiation SNPs on broad risk-taking behaviour). The association observed between the smoking initiation PRS and multiple risk-taking behaviours and externalising disorders in childhood suggests that the smoking initiation SNPs may be capturing a broader phenotype, such as risk-taking, which is not specific to smoking or nicotine, and thus this assumption may be violated. One approach which could be used to address this is Steiger filtering which can be used to exclude SNPs which explain the variance in the *outcome* over and above the variance in the *exposure* [10, 23]. The same approach can be applied in MR studies using smoking initiation PRS to remove SNPs which explain more variance in the negative control outcomes used in this study (or other phenotypes/proxies for risk-taking behaviour) than variance in smoking initiation. However, if the InSIDE assumption is perfectly violated (i.e., if the SNP effect on broad risk-taking causes smoking initiation), the smoking initiation PRS will be an invalid instrument using any MR method. At the very least, triangulating evidence across multiple MR methods (e.g., median weighted and mode based) would be advised in MR studies using smoking initiation PRS but, ideally, other causal inference methods should also be used.

There are a number of limitations of this study. First, the relatively low sample size – particularly when investigating associations with regular e-cigarette use and restricting to never smokers. Second, restricting analysis to never smokers could introduce collider bias [24]. We found that smoking initiation PRS were strongly associated with smoking initiation; if e-cigarette use causes young adults to smoke, then smoking status is a collider and conditioning on this variable (i.e., restricting analysis to never smokers) may inflate any association between smoking initiation PRS and e-cigarette use. Third, this cohort is not appropriate to directly study the gateway hypothesis as the young adults in ALSPAC were approximately 17 years old when e-cigarettes became widely available, and therefore were exposed to cigarettes earlier in their adolescence than e-cigarettes and had more opportunity to smoke than use e-cigarettes than later birth cohorts. Future research should explore this association in a larger sample of individuals with exposure to both cigarettes and e-cigarettes during adolescence. Fourth, the attrition rate in ALSPAC is considerable – only 2,905 of the 7,859 non-related participants of European ancestry with genetic data responded to the questions about vaping in the 24 year questionnaire – and missingness in this cohort has previously been associated with smoking initiation PRS [25]. Replicating the participation scores used by Taylor, Jones (25), we found that higher smoking initiation PRS were associated with participating in fewer ALSPAC questionnaires and clinics (change in participation per SD increase in smoking initiation PRS [*p* < 5×10^−8^ threshold] = -1.15, 95% CI -1.53 to -0.76). Furthermore, we found that those with higher smoking initiation PRS were less likely to have been included in the analysis of smoking initiation PRS and e-cigarette use due to attrition (OR_10-8_ per standard deviation of smoking initiation PRS = 0.87, 95% CI 0.83 to 0.91) so our estimates may be biased by selection and the association could be stronger than observed here. However, interpretation of any study including smoking initiation PRS will be difficult as the association between smoking initiation PRS and attrition could induce bias such as collider bias [26].

In conclusion, we find evidence to suggest there is a shared genetic aetiology between smoking and e-cigarette use but also with risky behaviour, SEP and externalising disorders in childhood. This suggests the PRS for smoking initiation is not specific to smoking or nicotine use but is capturing something much broader. Future research is needed to explore this in a population which has been exposed to both e-cigarettes and cigarettes in adolescence.

## Data Availability

The informed consent obtained from ALSPAC participants does not allow the data to be made freely available through any third party maintained public repository. However, data used for this submission can be made available on request to the ALSPAC Executive. The ALSPAC data management plan describes in detail the policy regarding data sharing, which is through a system of managed open access. Full instructions for applying for data access can be found here: http://www.bristol.ac.uk/alspac/researchers/access/. The ALSPAC study website contains details of all the data that are available (http://www.bristol.ac.uk/alspac/researchers/our-data/).

## Acknowledgements

We are extremely grateful to all the families who took part in this study, the midwives for their help in recruiting them, and the whole ALSPAC team, which includes interviewers, computer and laboratory technicians, clerical workers, research scientists, volunteers, managers, receptionists and nurses.

## Funding

This work was supported by the Medical Research Centre Integrative Epidemiology Unit at the University of Bristol [grant number MC_UU_0011/7]. The UK Medical Research Council and Wellcome (grant number 102215/2/13/2) and the University of Bristol provide core support for ALSPAC. This research was also supported by the NIHR Bristol Biomedical Research Centre at University Hospitals Bristol NHS Foundation Trust and the University of Bristol. The views expressed in this publication are those of the authors and not necessarily those of the NHS, the National Institute for Health Research or the Department of Health and Social Care. This publication is the work of the authors and JNK, REW, AET and MRM will serve as guarantors for the contents of this paper. A comprehensive list of grants funding is available on the ALSPAC website (http://www.bristol.ac.uk/alspac/external/documents/grant-acknowledgements.pdf). This research was specifically funded by CRUK (grant number C54841/A20491).

## Conflicts of interest

No conflicts of interest to declare.

## Notes

### Competing Interest Statement

The authors have declared no competing interest.

### Author Declarations

Ethics approval for the study was obtained from the ALSPAC Ethics and Law Committee and the Local Research Ethics Committees.

## References

1. Action on Smoking and Health. Use of e-cigarettes (vapourisers) among adults in Great Britain. 2019.

2. Hajek P, Phillips-Waller A, Przulj D, Pesola F, Myers Smith K, Bisal N, et al. A Randomized Trial of E-Cigarettes versus Nicotine-Replacement Therapy. N Engl J Med. 2019;380(7):629-37. Epub 2019/01/31. doi: 10.1056/NEJMoa1808779. PubMed PMID: 30699054.

3. Beard E, West R, Michie S, Brown J. Association between electronic cigarette use and changes in quit attempts, success of quit attempts, use of smoking cessation pharmacotherapy, and use of stop smoking services in England: time series analysis of population trends. 2016;354:i4645. doi: 10.1136/bmj.i4645 %J BMJ.

4. Khouja JN, Suddell SF, Peters SE, Taylor AE, Munafò MR. Is e-cigarette use in non-smoking young adults associated with later smoking? A systematic review and meta-analysis. 2020:tobaccocontrol-2019-055433. doi: 10.1136/tobaccocontrol-2019-055433 %J Tobacco Control.

5. Allegrini AG, Verweij KJH, Abdellaoui A, Treur JL, Hottenga JJ, Willemsen G, et al. Genetic Vulnerability for Smoking and Cannabis Use: Associations With E-Cigarette and Water Pipe Use. Nicotine Tob Res. 2019;21(6):723-30. Epub 2018/07/28. doi: 10.1093/ntr/nty150. PubMed PMID: 30053134.

6. Davey Smith G, Hemani G. Mendelian randomization: genetic anchors for causal inference in epidemiological studies. Hum Mol Genet. 2014;23(R1):R89-98. Epub 2014/07/30. doi: 10.1093/hmg/ddu328. PubMed PMID: 25064373; PubMed Central PMCID: PMCPMC4170722.

7. Heath AC, Madden PA, Slutske WS, Martin NG. Personality and the inheritance of smoking behavior: a genetic perspective. Behav Genet. 1995;25(2):103-17. Epub 1995/03/01. doi: 10.1007/bf02196921. PubMed PMID: 7733853.

8. Liu MZ, Jiang Y, Wedow R, Li Y, Brazel DM, Chen F, et al. Association studies of up to 1.2 million individuals yield new insights into the genetic etiology of tobacco and alcohol use. Nature Genetics. 2019;51(2):237-+. doi: 10.1038/s41588-018-0307-5. PubMed PMID: WOS:000457314300010.

9. Davey Smith G, Phillips AN, Neaton JD. Smoking as Independent Risk Factor for Suicide - Illustration of an Artifact from Observational Epidemiology. Lancet. 1992;340(8821):709-12. PubMed PMID: WOS:A1992JN78000014.

10. Hemani G, Bowden J, Davey Smith G. Evaluating the potential role of pleiotropy in Mendelian randomization studies. Human molecular genetics. 2018;27(R2):R195–R208. doi: 10.1093/hmg/ddy163. PubMed PMID: 29771313.

11. Fraser A, Macdonald-Wallis C, Tilling K, Boyd A, Golding J, Davey Smith G, et al. Cohort Profile: the Avon Longitudinal Study of Parents and Children: ALSPAC mothers cohort. Int J Epidemiol. 2013;42(1):97-110. Epub 2012/04/18. doi: 10.1093/ije/dys066. PubMed PMID: 22507742; PubMed Central PMCID: PMCPMC3600619.

12. Boyd A, Golding J, Macleod J, Lawlor DA, Fraser A, Henderson J, et al. Cohort Profile: The ‘Children of the 90s’-the index offspring of the Avon Longitudinal Study of Parents and Children. International Journal of Epidemiology. 2013;42(1):111–27. doi: 10.1093/ije/dys064. PubMed PMID: WOS:000316699300012.

13. Northstone K, Lewcock M, Groom A, Boyd A, Macleod J, Timpson N, et al. The Avon Longitudinal Study of Parents and Children (ALSPAC): an update on the enrolled sample of index children in 2019. Wellcome Open Res. 2019;4:51. Epub 2019/04/26. doi: 10.12688/wellcomeopenres.15132.1. PubMed PMID: 31020050; PubMed Central PMCID: PMCPMC6464058.

14. Harris PA, Taylor R, Thielke R, Payne J, Gonzalez N, Conde JG. Research electronic data capture (REDCap)--a metadata-driven methodology and workflow process for providing translational research informatics support. J Biomed Inform. 2009;42(2):377-81. Epub 2008/10/22. doi: 10.1016/j.jbi.2008.08.010. PubMed PMID: 18929686; PubMed Central PMCID: PMCPMC2700030.

15. Euesden J, Lewis CM, O’Reilly PF. PRSice: Polygenic Risk Score software. Bioinformatics. 2015;31(9):1466-8. Epub 2015/01/01. doi: 10.1093/bioinformatics/btu848. PubMed PMID: 25550326; PubMed Central PMCID: PMCPMC4410663.

16. StataCorp. Stata Statistical Software. 15.1 ed. College Station, TX: StataCorp LLC; 2017.

17. Khouja JN, Taylor AE, Munafò MR. Associations between reasons for vaping and current vaping and smoking status: Evidence from a UK based cohort. 2019:19006007. doi: 10.1101/19006007 %J medRxiv.

18. Action on Smoking and Health. Use of e-cigarettes (vapourisers) among adults in Great Britain. 2017.

19. Khouja JN, Munafò MR, Relton CL, Taylor AE, Gage SH, Richmond RC. Investigating the added value of biomarkers compared with self-reported smoking in predicting future e-cigarette use: Evidence from a longitudinal UK cohort study. 2019:19006247. doi: 10.1101/19006247 %J medRxiv.

20. Kim S, Selya AS. The Relationship Between Electronic Cigarette Use and Conventional Cigarette Smoking Is Largely Attributable to Shared Risk Factors. Nicotine & Tobacco Research. 2019. doi: 10.1093/ntr/ntz157.

21. Treur JL, Demontis D, Smith GD, Sallis H, Richardson TG, Wiers RW, et al. Investigating causality between liability to ADHD and substance use, and liability to substance use and ADHD risk, using Mendelian randomization. Addiction Biology. 2019. doi: 10.1111/adb.12849. PubMed PMID: WOS:000496650500001.

22. Davey Smith G, Ebrahim S. ‘Mendelian randomization’: can genetic epidemiology contribute to understanding environmental determinants of disease?*. International Journal of Epidemiology. 2003;32(1):1–22. doi: 10.1093/ije/dyg070.

23. Hemani G, Bowden J, Haycock P, Zheng J, Davis O, Flach P, et al. Automating Mendelian randomization through machine learning to construct a putative causal map of the human phenome. bioRxiv. 2017:173682. doi: 10.1101/173682.

24. Cole SR, Platt RW, Schisterman EF, Chu HT, Westreich D, Richardson D, et al. Illustrating bias due to conditioning on a collider. International Journal of Epidemiology. 2010;39(2):417–20. doi: 10.1093/ije/dyp334. PubMed PMID: WOS:000276303800019.

25. Taylor AE, Jones HJ, Sallis H, Euesden J, Stergiakouli E, Davies NM, et al. Exploring the association of genetic factors with participation in the Avon Longitudinal Study of Parents and Children. International Journal of Epidemiology. 2018;47(4):1207–16. doi: 10.1093/ije/dyy060 %J International Journal of Epidemiology.

26. Munafò MR, Tilling K, Taylor AE, Evans DM, Davey Smith G. Collider scope: when selection bias can substantially influence observed associations. Int J Epidemiol. 2018;47(1):226-35. Epub 2017/10/19. doi: 10.1093/ije/dyx206. PubMed PMID: 29040562; PubMed Central PMCID: PMCPMC5837306.

